# Risk perception of COVID-19 and its socioeconomic correlates in the United States: A social media analysis

**DOI:** 10.1101/2021.01.27.21250654

**Authors:** Shan Qiao, Zhenlong Li, Chen Liang, Xiaoming Li, Caroline Ann Rudisill

## Abstract

Social media analysis provides a new approach to monitoring and understanding risk perceptions regarding COVID-19 over time. Our current understandings of risk perceptions regarding COVID-19 do not disentangle the three dimensions of risk perceptions (perceived susceptibility, perceived severity, and negative emotion) over a long enough timeframe to cover different pandemic phases. The impact of social determinants of health factors on COVID-19-related risk perceptions over time is also not clear. To address these two knowledge gaps, we extracted tweets regarding COVID-19-related risk perceptions and developed index indicators for three dimensions of risk perceptions based on over 297 million geotagged tweets posted by over 3.5 million Twitter users from January to October 2020 in the United States. We also examined correlations between index indicator scores and county-level social determinants of health factors. The three domains of risk perceptions demonstrate different trajectories. Perceived severity kept climbing throughout the whole study period. Perceived susceptibility and negative emotion declined and remained stable at a lower level after peaking on March 11 (WHO named COVID-19 a global pandemic). Attention on risk perceptions was not exactly in accordance with epidemic trends of COVID-19 (cases, deaths). Users from socioeconomically vulnerable counties showed lower attention on perceived severity and susceptibility of COVID-19 than those from wealthier counties. Examination of trends in tweets regarding the multiple domains of risk perceptions throughout stages of the COVID-19 pandemic can help policy makers frame in-time, tailored, and appropriate responses to prevent viral spread and encourage preventive behavior uptake in United States.

## 1. INTRODUCTION

The coronavirus disease 2019 (COVID-19) pandemic caused by SARS-CoV-2 has spread across the world bringing severe morbidity and mortality and straining health care systems. By the end of 2020, the cumulative number of coronavirus cases globally exceeded 80 million and over 1,753,000 people had died of COVID-19. The United States has led the world in COVID-19 fatalities (over 400,000 as of January 20, 2021) (Johns Hopkins University and Medicine, 2020). The pandemic has profoundly and adversely impacted various aspects of society from health systems and economic growth to individuals’ day-to-day life, health and wellbeing.

Individuals have had to learn how to cope and adjust their life and expectations as the COVID-19 threat continuously evolves. The ebbs and flows of case rates in one’s individual environment as well as resulting movement restrictions, school closures and other changes to everyday life require constant adaption and assimilation of new information. These changes can also bring emotional side effects such as worry and anxiety. As part of this process of informational assimilation, risk perceptions would be expected to update over time.

Risk perceptions regarding COVID-19 also likely vary not just according to time but also across populations and socioeconomic contexts. Risk perception refers to individuals’ subjective assessments and appraisals about the probability of experiencing harms or hazards such as injury, illness, and death. According to a recent review on risk perceptions and risk characteristics, (Paek & Hove, 2017) risk perceptions are often composed of two main domains: the cognitive domain, which is about how much people understand risks (e.g., perceived susceptibility, perceived severity), and the emotional domain, which captures how people feel about risks (e.g., fear, dread). Many health behavior theories such as the Health Belief Model (HBM), Protection Motivation Theory (PMT), and the Risk Perception Attitude (RPA) framework emphasize the rational and cognitive aspects of risk perceptions. (Janz & Becker, 1984; Rimal & Real, 2003)

Slovic and colleagues highlighted the tendency to respond based on current emotions when understanding and making judgments about risks. For example, feeling intense dread may make people evaluate a risk as more threatening and prevalent. (Slovic, Finucane, Peters, & MacGregor, 2007) Some hypotheses predict that emotional reactions to risks (e.g., fear about the disease) could be independent of cognitive judgment and act as even stronger determinants of individual behaviors (Loewenstein, Weber, Hsee, & Welch, 2001). The degree to which the cognitive and emotional domains drive overall risk perceptions likely depends on the type of risk. The psychological approach suggests that risks have classifications such as an “unknown risk” (the risk that is new and unknown to science) and “dread risk” (the risk elicits visceral feelings of terror, uncontrollable, catastrophe, inequality, and uncontrolled), with the latter being influenced by emotions. (Slovic, 1987; Slovic, Fischhoff, & Lichtenstein, 1982; Visschers & Siegrist, 2018; Weber, 2017) The classification of a risk as unknown, dread, novel or experienced might then better predict updating of risk perceptions and behavioral response (Kousky, Pratt, & Zeckhauser, 2010)

Risk perceptions of a specific disease are generally expected to influence people’s health behaviors including preventive measures and vaccine uptake (e.g., H1N1 pandemic).(Agüero, Adell, Giménez, Medina, & Continente, 2011; Prati, Pietrantoni, & Zani, 2011; Rubin, Amlôt, Page, & Wessely, 2009; Rudisill, 2013; van der Weerd, Timmermans, Beaujean, Oudhoff, & van Steenbergen, 2011) Generally, a higher level of perceived susceptibility, perceived severity, and emotions (fear) are related to uptake of protective behaviors and willingness to vaccinate. Some empirical studies on risk perception and health-related behavior including intentions to vaccine have been conducted since the COVID-19 outbreak in the globe. (Detoc et al., 2020; Fisher et al., 2020) For example, one survey conducted among a nationally representative sample of 6,684 people in the United States during March 2020 reported large disagreement regarding their risks of COIVD-19 infection and infection fatality, which is to be expected given the survey timing. However, people who perceived higher risks were more likely to implement protective behaviors such as handwashing and social distancing.(de Bruin & Bennett, 2020) Several studies conducted among general populations in Europe suggested that perceived susceptibility and perceived severity are predictors of intention to vaccinate. (Dror et al., 2020; Graffigna, Palamenghi, Boccia, & Barello, 2020) One study in France showed that a higher level of fear about COVID-19 was related to higher vaccine acceptance. (Detoc et al., 2020) A recent study among 1,062 college students in the Southern United States found that perceived severity was positively associated with vaccine acceptance. (Qiao, Tam, & Li, 2020) A study of 253 young adults in Poland suggested that being worried about health was positively associated with willingness to obey strict hygiene and social distancing restrictions. (Sobkow, Zaleskiewicz, Petrova, Garcia-Retamero, & Traczyk, 2020) Therefore, we see evidence of relationships between perceived severity and behavioral response and vaccination acceptability. However, work that has examined the difference in perceptions about COVID-19 risk for oneself versus others and the impact on intentions to vaccinate from the UK has found that perceptions of risk for others are more predictive of behavioral response (Sherman et al., 2020)

Given that risk perceptions are important precursors to health-related behaviors for either dealing with or preventing risk, many models and theories have been developed to identify the factors at various socioecological levels that may influence risk perceptions. At the structural level, social determinant of health factors may influence people’s health beliefs, health literacy and self-efficacy as well as shape their experience of risk. In the context of COVID-19, the factors associated with risk perceptions have been identified as age, gender, pre-existing health conditions,(Alschuler, Roberts, Herring, & Ehde, 2020; Asefa, Qanche, Hailemariam, Dhuguma, & Nigussie, 2020; Ding et al., 2020; Guastafierro et al., 2021; Ning et al., 2020) knowledge of COVID-19, working at or outside of home, (Mansilla Dominguez et al., 2020) experience with the virus within family and social networks, and some psychological factors such as anxiety and distress.(Alschuler et al., 2020; Orte, Sanchez-Prieto, Dominguez, & Barrientos-Baez, 2020) Political partisanship and trust in governments (e.g. local, national) may also shape the risk perceptions.(Barrios & Hochberg, 2020; Ye & Lyu, 2020) Socioeconomic (SES) factors such as residence and economic status have been found to influence self-efficacy of responding to risk. Jahangiry and colleagues discovered that living in rural areas and having good economic status was positively related to higher perceived efficacy (i.e., self-efficacy and response efficacy) among the Iranian general population (online survey of 3727 individuals). (Jahangiry et al., 2020)

Social media has been playing an important role in reflecting and reshaping people’s perceptions of COVID-19 risk. Social media platforms such as Twitter have become a critical source for information exchange and an outlet for users to express their opinions and concerns and share experiences and feelings about the pandemic. Social media data are characterized by being real-time and anonymous (to some extent). The nature of social networking often provides rapid response and peer feedback. In addition, these data are high volume (e.g., approximately 500 million tweets per day on Twitter), (Krikorian, 2013) provide large population coverage (e.g., 67% of 18-29 years old adults in the US are likely to use *Instagram*), (Pew Research Center, 2019) offer a variety of data formats (e.g., text, images, video, geospatial data), and are highly available and affordable for analysis.

Social media analysis, especially tweet analysis, provides a new approach to monitoring and exploring risk perceptions in a long period based on large dataset. Given the constantly changing pandemic in each individual’s environment and on a global scale, the dynamic nature of social media data provides a rapid window into perceptions of risks. Robust social media analysis can be used to inform health promotion strategies including vaccination uptake. Chandrasekaran and colleagues conducted text mining of COVID-19-related English tweets from January to May 2020 to group the main topics and uncover the key trends by examining sentiment (positive or negative) scores. (Chandrasekaran, Mehta, Valkunde, & Moustakas, 2020) Dyer and Kolic developed indicators of public risk perception based on emotion and attention presented in tweets from 12 countries between March and June 2020. Twitter users showed differential sensitivity by country to national COVID-19 death rates.(Dyer & Kolic, 2020)

Existing work does not, however, address a number of crucial aspects that are important for using such a rapid response mechanism to understand population-level COVID-19 pandemic responses. First, existing studies on risk perceptions do not disentangle the three dimensions of risk perceptions (perceived susceptibility, perceived severity and negative emotion) over a long enough timeframe to investigate risk perceptions across different phases of the pandemic. A longer timeframe allows for a better understanding of the nuances in risk perception changes as actual risk and experience with COVID-19 vary over time. Second, existing work does not connect social determinants of health factors (via data such as county-level US Census data) with social media data to understand these factors that are known to impact risk perceptions.

To address these knowledge gaps, the current study aims to 1) demonstrate the trajectories of perceived susceptibility, perceived severity, and negative emotion since the start of the COVID-19 outbreak in the United States based on tweets from January to October 2020; 2) illustrate the degree to which these three trajectories are in accordance with epidemiological COVID-19 trends (e.g., daily new cases and daily new death); and 3) examine which social determinants of health factors correlate with the three domains of risk perceptions based on county level SES data in the United States.

## 2. DATA AND METHODS

### 2.1. Data Sources

#### 2.1.1. Geotagged Twitter data

We collected more than 305 million (N= 305,150,205) tweets posted by over 3.5 million (N= 3,539,309) Twitter users from January 1, 2020 to October 20, 2020 using the free public Twitter streaming application programming interface. (Twitter, 2020) All tweets were geotagged (embedded with a geolocation) within the continental United States. Following our previous work, (Martin, Cutter, Li, Emrich, & Mitchell, 2020) we filtered out tweets automatically posted by bots such as weather reports and job offers by checking from which application a tweet was posted (the source of a tweet). After data cleaning, 297,354,262 geotagged tweets posted by over 3,520,692 Twitter users were left for further analysis to identify tweets relevant to the research aims.

#### 2.1.2. COVID-19 epidemic data and key events

U.S. national-level and county-level daily accumulated COVID-19 confirmed cases and deaths were downloaded from the New York Times’ GitHub data repository. (New York Times, 2020)The national-level daily new cases and deaths were derived from accumulated data. A list of key events since the COVID-19 outbreak were extracted from news reports and government announcements (see Supplement Table S1).

#### 2.1.3. Socioeconomic and demographic data

The U.S. county-level socioeconomic (SES) and demographic (race/ethics) variables were extracted from 2014-2018 American Community Survey (ACS) 5-Year estimates data.(U.S. Census, 2019) The SES variables include Gini coefficient, median household income, percent of unemployed, percent of having no health insurance, percent of living in poverty, and percent of having education less than high school. The race/ethnicity variables include percent of Black or African American, percent of White, percent of Hispanic or Latino, and percent of Asian. Lastly, county-level population density was also derived from the 2014-2018 ACS data.

### 2.2. Keywords Identification for Risk Perceptions

Three categories of risk perception keywords were identified based on literature, including perceived susceptibility, perceived severity, and negative emotional dimension (Table 1). Perceived susceptibility captures people’s subjective beliefs about how vulnerable and susceptible they are to a disease or other health risk, that is, likelihood or probability of getting the disease. Perceived severity captures how serious people believe a health risk to be and whether it will have adverse physical consequences such as death, disability, and pain, and adverse social consequences such as ostracism, stigma, and shame. The emotional dimension depicts how people feel about the risks, such as fear, outrage, dread, etc.

**Table 1.** Identified keywords for the three domains of COVID-19 risk perceptions.

| **Perceived Susceptibility**<br>**(CHV Ontology ID)** | **Perceived Severity**<br>**(CHV Ontology ID)** | **Negative Emotion**<br>**(CHV Ontology ID)** |
| --- | --- | --- |
| Vulnerable/vulnerate | Die | Worse/worthen/worthening |
| Risk/risky | Dead/death | Worthened/worst |
| Unsafe/not safe (ochv#37555) | Lethal | Dread |
| Suspect | Fatal | Fear/feared/fearful/fearing |
| Doubt/dubious | Pain/painful | (ochv#37463) |
| Hesitate/hesitating | (ochv#9185) | Scare/scared/scaring |
| Danger/dangerous | Isolate | (ochv#51823) |
| Unsure | Judge | Outrage |
| Believe/believed | Shame/shameful | Nervous |
| Undoubted/undoubting | Suffer/suffering/suffered | Panic |
| Confused/confusing/confusion | Paralyzed | Terrify/terrified/terrifying |
| Immune /immunity | Restricted | Worry/worried |
| High risk/ high-risk |  | Anxious/anxiety |
| At risk/ at-risk |  | Stress/stressed |
| Avoid |  | Distrust |
| Cancel |  |  |
| Postpone |  |  |

Note: a complete ID in CHV ontology is http://sbmi.uth.tmc.edu/ontology/[identical ID of a concept]

Individual risk perception keywords were identified by two researchers (SQ and CR) in review of the Linguistic Inquiry and Word Count (LIWC), a closed vocabulary of cognitive and emotional terms used by laypersons. (Tausczik & Pennebaker, 2010) Keywords that were used during consumer health communications were mapped onto the Ontology of Consumer Health Vocabulary (Amith, Cui, Roberts, Xu, & Tao, 2019)(Table 1), a formal and interoperable semantic web ontology that was developed based on the Consumer Health Vocabulary (CHV). (Zeng & Tse, 2006) Therefore, these identified keywords were validated by human experts, standardized by LIWC and CHV, and enhanced in term of generalizability as part of them could be semantically linked to existing medical/healthcare vocabularies as identified by the Uniformed Medical Language System (UMLS).(Bodenreider, 2004)

### 2.3. Risk Perception Index

We defined the risk perception index (RPI) for a specific risk perception domain as the proportion of Twitter users who posted domain-specific risk perception tweets among all Twitter users who posted COVID-19 related tweets (Equation 1). The COVID-19 related tweets were extracted using the following keywords: coronavirus, covid-19, covid19, pandemic, epidemic, and virus.

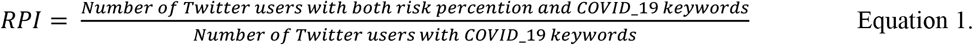

At the national level, a daily RPI was computed for each domain of risk perceptions from January 1 to October 20, 2020. The daily RPI was used for temporal analysis to demonstrate the trajectories of each risk perception domain since the COVID-19 outbreak in the United States. At the county level, an accumulated RPI for each county of each risk perception domain was computed by using the aggregated Twitter data within the study period. When computing the county level index, we only included tweets that were geotagged with a spatial resolution at or finer than the city level so that we were able to associate each tweet with a county. The accumulated RPI was used for statistical analysis at the county level to examine how social determinants of health factors correlate with the three domains of risk perceptions.

### 2.4. Data Analysis

#### 2.4.1. Temporal trend analysis at the national level

The daily RPI for each of three risk perception domains (perceived susceptibility, perceived severity, and negative emotion) were computed and plotted as time series at the national level. This allows us to depict their trajectories since the COVID-19 outbreak in the United States based on tweets from January to October. To illustrate the degree to which these three trajectories were in accordance with the trend of COVID-19 pandemic, COVID-19 epidemic data (number of daily new cases and daily new deaths) were overlayed and visually associated with the risk perception indices. Lastly, a list of selected key events since the COVID-19 outbreak were also connected to the trend lines to examine how these key events affected posted tweets related to risk perceptions.

#### 2.4.2. Statistical analysis at the county level

To examine how social determinants of health factors and demographic variables correlate with attention to the three domains of risk perceptions, correlation analysis was performed between the county-level risk perception indices and county-level SES variables (Gini coefficient, median household income, percentage of being unemployed, percentage of having no health insurance, percentage of living in poverty, and percentage of having education less than high school) and race/ethics variables (percentage of Black or African American, White, Hispanic or Latino, and Asian). To reduce uncertainty, counties with less than 100 Twitter users posted COVID-19 related tweets were removed, resulting in 754 counties included in the statistical analysis.

## 3. RESULTS

### 3.1. Temporal Trend of Twitter-derived Risk Perceptions

Figure 1 illustrates the trajectories for the three domains of risk perceptions from January 1 to October 20, 2020 covering three phases of the COVID-19 pandemic evolving process in the United States: Phase I: initial outbreak to global pandemic (January to March); Phase II: lockdown as a response to this public emergency (April to May); and Phase III: reopen and surge in cases (June to October).

**Figure 1.**
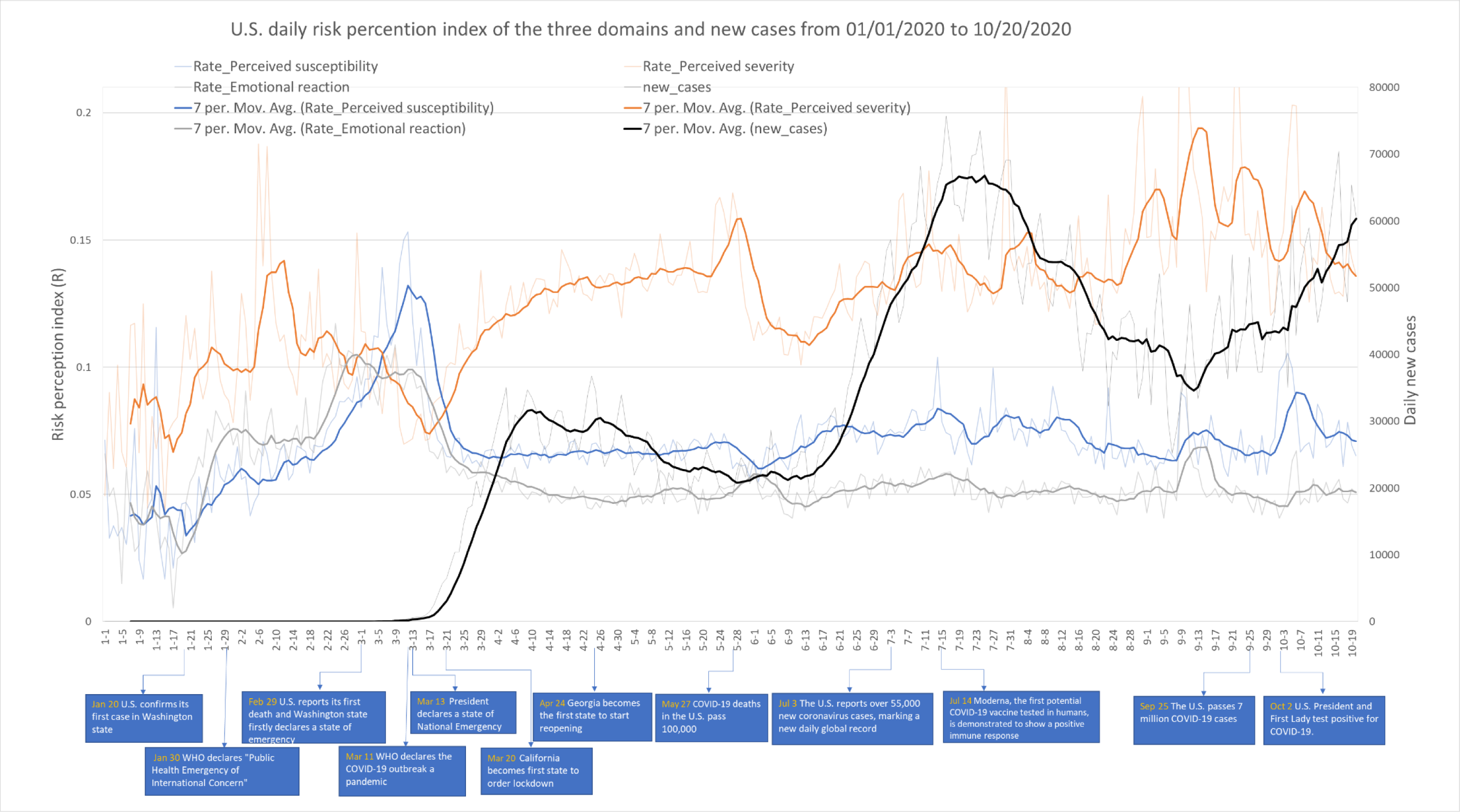
Trajectories for changing trends of three risk perception domains and COVID-19 daily new cases (with big events and milestones for multiple phases of the pandemic)

The trajectories of perceived susceptibility and negative emotion were in accordance with each other displaying a great difference to the trajectory of perceived severity. The perceived severity index score was generally higher than the ones of perceived susceptibility and negative emotion throughout the whole study period. In addition, the trajectory of perceived severity kept increasing over all phases, while the other two trajectories peaked after March 11 when the WHO declared COVID-19 a global pandemic and then declined a little bit and remained stable in Phase II and III. In Phase II, the index score of perceived severity spiked up on the week of May 28 when COVID-19 deaths in the U.S. passed 100,000. In Phase III, the trajectory of perceived severity climbed to a higher plateau in September when COVID-19 cases in the U.S. were over 7 million. A spike in the index score of negative emotion appeared in Phase III with the news that the U.S. President and First Lady tested positive for COVID-19.

Comparison between the trajectories of COVID epidemiological indicators (daily new cases and daily new deaths) suggests that the trends in the pandemic’s epidemiological status were not exactly aligned with changes in perceptions regarding the COVID epidemic (See Figure 1 and 2). The temporal trend of Twitter-derived perceived severity lagged one month or so behind the real-time change of daily new cases.

**Figure 2.**
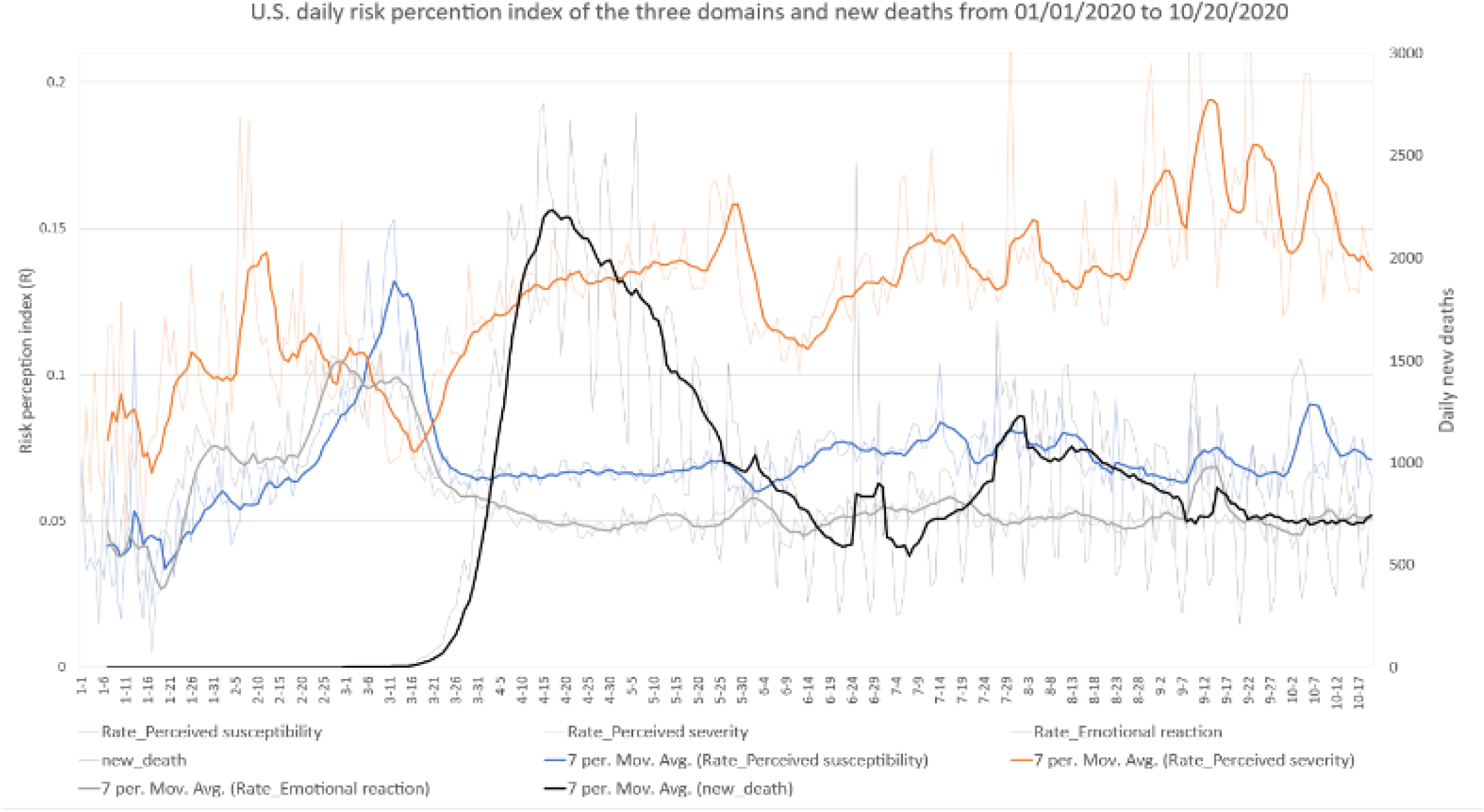
Trajectories for changing trends of three risk perception domains and COVID-19 daily new deaths.

### 3.2. Correlations between Risk Perceptions and Socioeconomic and Demographic Factors

Table 2 shows the correlation results between county-level socioeconomic and demographic variables (e.g., SES, race, population density) and the index scores of the three risk perception domains. In general, low SES levels were correlated with low index scores of perceived severity and susceptibility. For example, high percentages of having no health insurance and education attainment less than high school were significantly related to a low index score of perceived susceptibility and severity of COVID-19 (p<.01). In addition, the index score of perceived severity was positively correlated with median household income (p<.05) and negatively with percentage of living in poverty (p<.01). When examining demographic correlates, we found that a high percentage of Black people in a county was related to a low index score of perceived severity, susceptibility and negative emotion regarding COVID-19, while a high percentage of White people in a county was related to a higher index score of all three risk perception domains. No significant correlations were observed between the percentage of Hispanic/Latino or Asian in a county and the index score of any risk perception domain. There was no significant correlation between the risk perception index score and county-level population density.

**Table 2.** Correlation results between county-level SES and demographic variables and the index score for three risk perception domains.

| **r** | **Perceived**<br>**susceptibility** | **Perceived**<br>**severity** | **Negative**<br>**emotion** |
| --- | --- | --- | --- |
| Gini coefficient | 0.0049 | **-0.0963\*\*** | -0.0483 |
| Median household income | -0.0131 | **0.0936\*** | -0.0322 |
| Percentage of being unemployed | 0.0008 | 0.0168 | 0.0518 |
| Percentage of no health insurance | **-0.1775\*\*** | **-0.1838\*\*** | -0.0524 |
| Percentage of living in poverty | 0.0261 | **-0.1702\*\*** | -0.0127 |
| Percentage of less high school | **-0.1144\*\*** | **-0.1515\*\*** | -0.0095 |
| Percentage of African American | **-0.1931\*\*** | **-0.2183\*\*** | **-0.1940\*\*** |
| Percentage of White | **0.1759\*\*** | **0.1703\*\*** | **0.1749\*\*** |
| Percentage of Hispanic/Latino | -0.0504 | -0.0292 | 0.0323 |
| Percentage of Asian | 0.0070 | 0.0396 | -0.0334 |
| Population density | -0.0106 | 0.0181 | -0.0247 |

\* $p < 0.05$ \*\* $p < 0.01$

## 4. DISCUSSIONS

Using over 297 million geotagged tweets posted by over 3.5 million Twitter users from January to October 2020 in the United States, we extracted tweets regarding risk perceptions of COVID-19 and developed index indicators for its three domains (i.e., perceived severity, susceptibility, and emotion). We demonstrated and compared the trajectories of the three domains with the COVID-19 epidemic trend during an almost 10-month timeframe covering different phases of the pandemic’s evolution. To the best of our knowledge, this work is one of the first efforts to investigate how county-level socioeconomic and demographic factors correlate with the three domains of COVID-19 risk perceptions based on social media analysis. It is also the first work using social media data that captures the COVID-19 pandemic from its start to late fall 2020.

The three domains of risk perceptions demonstrate different trajectories. Generally, Twitter users were more concerned about the severity of COVID-19 rather than its perceived susceptibility or demonstrating negative emotion as index score of severity kept climbing throughout the whole study period. Conversely, the index score of perceived susceptibility and negative emotion declined and remained stable at a lower level after peaks on March 11 when COVID-19 was named a global pandemic by the WHO. Perceived susceptibility and negative emotion did, however, have very similar patterns both in level and trajectory.

Importantly and in contrast to assumptions that actual risk may play a factor in perceived risk, attentions/discussion on perceptions of COVID-19 risk were not exactly in accordance with trends in COVID-19 epidemic indicators (e.g., daily new cases, daily new deaths). However, they do appear to be triggered by big news or events (e.g., President and First Lady Trump’s COVID-19 diagnoses). Finally, our study suggests that social determinants of health factors such as income, race, education level, poverty and health insurance were correlated with discussions regarding risk perceptions. Users from socioeconomically vulnerable counties showed lower attention on perceived severity and susceptibility of COVID-19.

Our findings demonstrate that perception and understanding of risks regarding a new public health threat is complicated and people’s attention to multiple domains may be evolving differently. The climbing trajectory of perceived severity implies the general population’s growing awareness of this new virus as related scientific discoveries emerge; however, this does not appear related to actual cases or deaths. The increased attention to severity over time might not contribute to more discussion about perceived susceptibility and thus might not translate into associated behavioral response. A survey of 1,591 people in the United States over the first week of the pandemic found growing awareness of general COVID-19 risk but underestimated infection susceptibility relative to that of the average person in the US. (Wise, Zbozinek, Michelini, Hagan, & Mobbs, 2020) Optimism bias may be one reason as has been found in previous pandemics (e.g. H1N1) although this was not found to impact behavioral response. (Rudisill, 2013)

Another explanation for the relatively low index score of perceived susceptibility may be increasing self-efficacy in knowledge and skills regarding public prevention. In Phase I, there was so much uncertainty about the new virus and how it spreads even among scientists and public health experts that no one could expect the public to have certainty. In Phases II and III, however, people understood more about how to protect themselves from infection, developed more confidence in controlling their own pandemic-related risks, and increased self-efficacy in conducting protective behaviors based on more concrete advice on prevention strategies. They also had more personal experience living in this environment and increasingly adjusted to new norm.

The index score of negative emotion remained stable at a lower level across the three phases after a peak when the WHO declared COVID-19 a global pandemic. There are several potential explanations. First, many people started feeling fatigue due to continual exposure to COVID-19 related reporting. As the pandemic evolved, they might become numb about the news and reduce their frequency in posting or reposting on Twitter about negative feelings. Second, the low level of the index score for negative emotions was consistent with that of perceived susceptibility. The low level of attention to perceived susceptibility implies a feeling that “at least I am safe from the COVID-19 risk”, which might buffer negative emotion such as anxiety or panic caused by COVID-19 pandemic. Third, COVID-19, certainly in the beginning of 2020 and over 2020 has been an “unknown risk” rather than “dread risk” for the public. According to existing psychological theories on risk perception, “unknown risk” is not as closely associated with emotions as “dread risk” is. To some extent, the number of COVID cases does not always evoke strong emotions or feelings. Many people have difficulties in understanding numerical information related to risk. (Cokely, Galesic, Schulz, Ghazal, & Garcia-Retamero, 2012) Finally, positive coping strategies, resilience, and social support might help people to bounce back from various negative emotions during the period of lockdown in Phase I. People might develop positive emotions and self-efficacy in coping with this public health threat. For example, sentiment analysis based on COIVD-19 related tweets from January to May 2020 suggested a reversal of sentiments from negative to positive for topics such as public prevention, government response, impact on healthcare industry, and COVID-19 treatment and recovery (Chandrasekaran et al., 2020)

It is notable that the trajectories of risk perceptions were not in accordance with those of COVID-19 epidemiological indicators. However, the trajectory of perceived susceptibility seemed to show a “lag” behind the trend of daily new cases of COVID-19. This finding suggests that perception of risks is a procedure of accessing, extracting, and digesting information about this new virus. It could be a “learning process” and also a “re-constructing” of attitudes and health beliefs. Therefore, it is not surprising that people’s attention and discussion of the risks may be delayed upon receipt of new information.

Our study suggests that low SES status in one’s county where tweeting was correlated with lower attention to perceived severity and susceptibility of COVID-19. Poverty might limit people’s in-time access to accurate information on COVID-19. Low educational attainment could lead to low health literacy, which increased difficulties in understanding health information.(Friis, Lasgaard, Rowlands, Osborne, & Maindal, 2016; Paakkari & Okan, 2020) African American people, influenced by their cultural contexts and health beliefs and a history of distrust in health system, (Boulware, Cooper, Ratner, LaVeist, & Powe, 2016) might underestimate their risks of infection. (Eiser & Ellis, 2007; Paakkari & Okan, 2020) For example, a study on COVID-19 related Tweets posted by African Americans (n=1,763) from Jan 21^st^ to May 3^rd^, 2020 reported that positive sentiments and optimism were uniquely observed in African American Twitter communities. The percentage of topics like Black strong (27.1%) and growing up Blacks (22.8%) was higher than COVID-19 prevention behaviors such as encouraging social distancing (9.4%) and masks (4.7%). (Odlum et al., 2020)

This study has several methodological limitations that require attention in interpreting and generalizing from findings. First, we need to be cautious about the representativeness of Twitter users. Twitter is not universally used in the United States, particularly among older and low-income populations. In addition, not all Twitter users share their geolocation information.

Therefore, those who geotag their tweets are not representative of the wider Twitter population. (Jiang, Li, & Ye, 2019) Second, we used index scores as proximal indicators to quantify people’s attention (relative frequency of tweet posts) to the three domains of the risk perceptions. We did not use existing validated measure instruments to assess the level of perceived severity, susceptibility, or negative emotions. Third, the keyword-based tweets retrieval method may miss a small number of relevant tweets that that did not include common language regarding risk perceptions. Specifically, keywords-based methods only capture tweets with an exact match of terms. Indirect mentions of risk-perception terms and subtle cues may be missed because human natural language is rich and dynamic. Text-mining methods, such as topic modeling will be needed as a supplement to further strengthen the understanding of people’s opinions. Fourth, in terms of emotion, we only examined negative emotional reaction to COVID-19 in the analysis. According to some theoretical frameworks, self-efficacy and resilience could be other domains of risk perceptions.(Jahangiry et al., 2020)Further studies are needed to investigate the trend of positive emotional reactions during different phases in the pandemic. Finally, limited by the scope of the current study, we were not able to elaborate the trajectories of risk perception index score during the COVID-19 pandemic at the county-level. Using an accumulated index score for each county limits the implications of temporal trends on how perceived risk perceptions interact with SES within a county.

Despite these limitations, the current study suggests that social media analysis integrated with geospatial data could be a promising tool for real-time monitoring of risk perceptions during a new public health threat. This work can then inform public health policy and intervention strategies in a dynamic way. A number of key policy implications emerge from this work. First, policy makers and public health professionals need to consider and monitor multiple domains of risk perception through different phases of the pandemic given that these domains demonstrate different trajectories across pandemic phases. Health communication and education interventions can be tailored for the public health emergency’s evolving stages. Public attention to perceived susceptibility and negative emotion may decrease as people feel they have learnt sufficiently about the new virus and been able to control it. This trend may lead to lower compliance with or even giving up on protective behaviors. This could contribute to another wave of COVID-19 outbreak as shown in United Kingdom and United States in the winter of 2020. Social marketing and health communication campaigns are needed to communicate to people with effective alerts and reminder messages when they have “prevention fatigue.” In addition, the next phase of behavior related to COVID-19 is vaccination. It is important to continue understanding the pulse of population perceptions of COVID-19 risk to target and design strategies to encourage vaccine uptake and reduce hesitancy.

Second, the “lagging effect” between the COVID-19 epidemic updates (e.g., daily new cases) and awareness of risk (e.g., perceived severity) has important implications in responding to a new public health threat. When people face a new virus for which they have no proximal risk to rely on for understanding and coping, there could be a large lag in developing and constructing their awareness and developing perceptions of the new risk. Furthermore, preventive action may be further delayed upon risk updating. Policy makers and health educators need to realize this lag and avoid underestimating the difficulties and challenges in spreading accurate information and promoting protective behaviors within a short-term. The similar lag may appear for COVID-19 vaccination campaigns and efforts to maintain other public health good practice.

Third, social determinant of health factors may influence the cognitive domain (perceived severity and susceptibility) rather than emotional domain of risk perceptions during the COVID-19 pandemic. Perceived severity and susceptibility are important predictors of compliance with preventive and protective behaviors during public health crises (such as in HIN1 and COVID-19). Low levels of attention to the cognitive domain may indicate low awareness of risk, which may further impede uptake of protective behaviors (e.g., social distancing, vaccine uptake). Extant literature has reported great health disparities during the COVID-19 epidemic regarding case rates and clinical outcomes in the United States.(Loomba et al., 2021; Okonkwo et al., 2020; Zhang & Schwartz, 2020) Given that people from counties with low SES may be more likely to be exposed to the virus because of their working conditions (e.g., they cannot work at home), low awareness of risk could double their vulnerability toward COVID-19. These groups should be prioritized for health education and promotion receiving tailored messages using understandable and culturally appropriate language.

In conclusion, examination of changing trends in tweets regarding multiple domains of risk perceptions throughout stages of the COVID-19 pandemic can help governments, policy makers and healthcare agencies frame in-time, tailored, and appropriate responses to prevent the pandemic’s spread in United States. Living in a county with relatively low SES status was correlated with a low level of attention to perceived severity and susceptibility of COVID-19.

Communities with low SES factors and high percentages of African Americans need to be prioritized in health communication campaigns and interventions. Key messages in social marketing and health promotion should be tailored in accordance with patterns of changing trends about risk perceptions across multiple phases of the pandemic. The lessons obtained in risk perceptions about COVID-19 spread thus far may inform effective intervention strategies for COVID-19 vaccine administration and other public health crisis in future.

## Data Availability

social media data is for public

## Acknowledgement

Research reported in this publication was supported by the National Institute of Allergy and Infectious Diseases of the National Institutes of Health under Award Number R01AI127203-4S1, National Science Foundation under Award Number NSF2029791.The project was also supported by the University of South Carolina Office of Vice President for Research COVID-19 grant (USCIP 80003673, PI: X Li). (135400-20-54176, PI: Z Li), and (115400-20-54086, PI: CA Rudisill). The content is solely the responsibility of the authors and does not necessarily represent the official views of the National Institutes of Health and University of South Carolina. We greatly appreciate Dr. Quan Zhang for his assistance in preparing figures and supplement document.

